# Towards a Buruli Ulcer Rapid Diagnostic Test that Targets Mycolactone

**DOI:** 10.1101/2024.01.23.24301643

**Authors:** Marina Siirin, Bijan Pedram, Maria J. Gonzalez-Moa, Louisa Warryn, Rie Yotsu, Jean M. Saunders, Aaron E. Saunders, Richard K. Baldwin, Jessica L. Porter, Timothy P. Stinear, Israel Cruz-Mata, Dziedzom Komi de Souza, Gerd Pluschke, Marco A. Biamonte

## Abstract

We report the development of a prototype rapid diagnostic test for Buruli ulcer, an ulcerative necrotizing skin disease caused by *M. ulcerans*. The test was designed to detect mycolactone, a metabolite unique to *M. ulcerans*. The chief technical challenge was to develop a simple workflow to extract trace amounts of mycolactone from wound exudates collected with a swab and, after a concentration step, to visualize the mycolactone by means of a lateral flow assay. This was achieved by utilizing a mouse monoclonal antibody specific for mycolactone and magnetic gold nanoshells. The latter are a novel class of reporter particles consisting of a ferrite core, a silica gel middle layer that serves to decrease the overall density of the nanoparticle and facilitate its resuspension in aqueous media, and an outer layer of gold, which provides a dark coloration through plasmon resonance effects. These nanoparticles, once conjugated to the anti-mycolactone antibody, enable the immunomagnetic concentration of the targeted analyte and its detection by lateral flow assay. The test procedure can be conducted within 2 hours with a magnetic rack and no powered instrumentation is required. The test can detect as little as 3.5–7 ng of mycolactone collected on a swab.

**Author summary:** Buruli ulcer is a neglected tropical disease that affects the poorest of the poorest in Africa. Even young people can harbor large ulcers, with raw flesh directly exposed. While antibiotic treatments exist, there are no simple methods to diagnose the disease. Here, we attempted to detect mycolactone, a small molecule produced by the bacteria that causes Buruli ulcer. One difficulty was the sample type to be used, as the open wounds are sampled with a swab. The question was how to extract mycolactone from the swab and then detect it by means of a rapid diagnostic test, without using sophisticated equipment. We tried to answer this question by combining monoclonal antibodies specific for mycolactone and novel magnetic nanoparticles to concentrate and visualize mycolactone with a rapid test.

## Introduction

Buruli ulcer (BU) is a progressive necrotizing skin infection caused by *Mycobacterium ulcerans.* The disease starts as a painless papule which can progress to a nodule and eventually degenerate into a large ulcer where the skin no longer covers the underlying tissue [1, 2]. Buruli ulcer has been reported in 34 countries [3], and is mostly found in rural areas of West Africa, though a recrudescence has been noted in Australia [4] where its transmission has been ascribed to possums and other animal reservoirs [5, 6]. In 2019, before the COVID-19 pandemic which affected active case finding campaigns, 2,271 new cases were recorded worldwide, and the incidence rate remained at the same level with 2,121 cases reported in 2022 [7]. The disease is believed to be vastly under-reported partly due to its presence in very remote communities hindering access to care, and also because of the lack of awareness, of reporting system, and of field-friendly diagnostic tools [8].

Buruli ulcer remains one of the neglected tropical diseases (NTDs) in need of improved diagnostic tests. With oral antibiotic treatment available [9], decentralizing diagnosis would facilitate strategies for early detection, timely treatment, and prevention of permanent disability. However, there are no diagnostics suitable for point-of-care applications. The well-established diagnostics are based on direct *M. ulcerans* pathogen confirmation by *in vitro* culture (definitive positive diagnosis), PCR, smear examination for acid-fast bacilli, or histopathology. These diagnostics require either a sophisticated laboratory or have low sensitivity. An isothermal amplification technique (LAMP assay) has been proposed as alternative to PCR and, while not commercial, is promising [10, 11]. A method based on thin layer chromatography has also been evaluated and works better with fine needle aspirates from nodules than with swabs from open wounds [12, 13].

Given the importance of securing new diagnostic tools for Buruli ulcer and to help guiding the development of new diagnostic tools, the World Health Organization (WHO) in consultation with different partners developed a preliminary Target Product Profile (TPP) in 2018 [14] which was revisited in 2022 [15]. The TPP served as the starting point for the efforts described herein. Of particular relevance to this work was the desire stated in the TPP to see a diagnostic test capable of detecting mycolactone. Mycolactone is a unique metabolite of *M. ulcerans* composed of a macrolide core substituted with a fatty-acid sidechain (MW = 743.02). Mycolactone has cytotoxic, immunosuppressive, and analgesic effects that induce the large debilitating skin lesions that develop when the infection is not treated [16]. It is further believed that because mycolactone production is related to the viability of the mycobacterium, a mycolactone test has the potential to be used not only for diagnosis, but also to assess response to treatment and cure.

Here, we report the development of a prototype rapid test for Buruli ulcer, formatted as a lateral flow assay capable of identifying mycolactone as the disease biomarker. The test was specifically designed to isolate trace amounts of mycolactone from wound exudates, and then analyze the concentrated mycolactone on a lateral flow assay with minimal sample processing.

## Materials and Methods

### Assay Principle

We devised a competitive lateral flow assay featuring two innovations. The first was the use of a mouse monoclonal antibody specific for mycolactone [17], which has previously shown value in an ELISA [18, 19]. The second innovation was the use of custom-made nanoparticles specifically designed to enable immunomagnetic concentration of the analyte mycolactone while being darkly colored and providing a visual signal when run on a lateral flow test. These nanoparticles, dubbed magnetic gold nanoshells (Mag-GNS) consist of a ferrite center, a silica gel middle layer, and a gold outer shell (**Figure 2**). The ferrite center allows for magnetic concentration, the silica gel layer encapsulates the ferrite and lowers the overall density of the nanoparticles allowing for their resuspension in aqueous media, and the gold shell surface absorbs light extremely efficiently due to plasmon resonance effects. This quantum physics phenomenon comes into play when metal nanoparticles reach the nm scale, in this case creating a dark brown-gray color, nearly black, a surprising hue for a gold particle. The gold shell is further functionalized with carboxylic acid groups for covalent conjugation to a detector antibody, and was custom made by nanoComposix, a Fortis LifeScience Company (San Diego, CA, USA).

**Figure 1.**
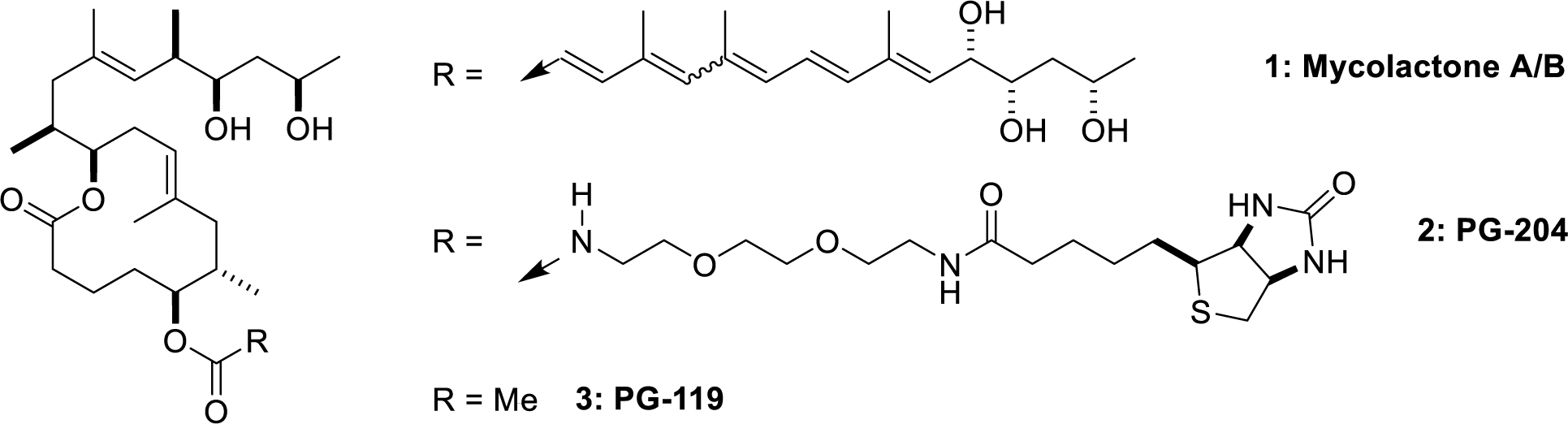
Structures of (1) mycolactone A/B isomer mixture, (2) biotinylated analog PG-204, and (3) acetyl analog PG-119.

**Figure 2.**
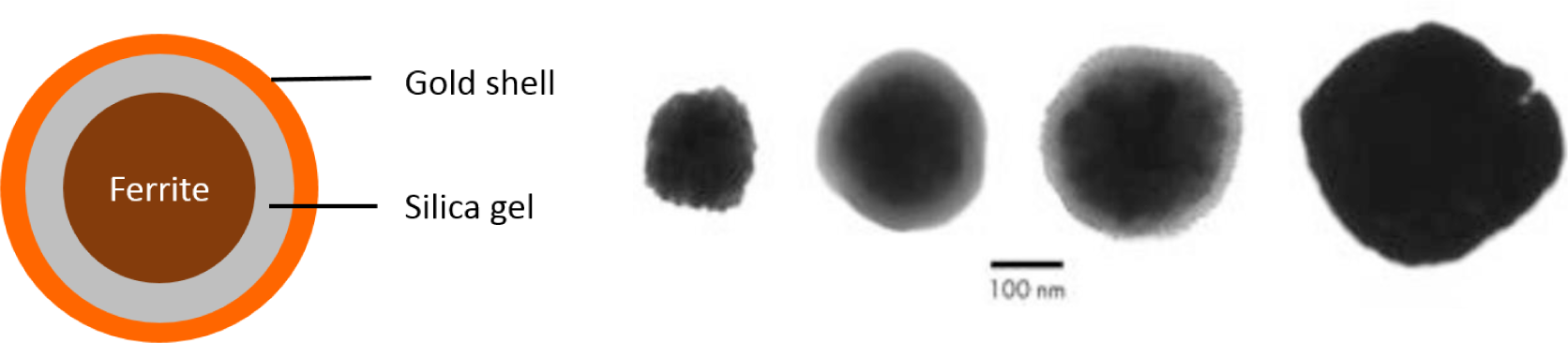
Magnetic gold nanoshells. Schematics and transmission electron microscopy views of the particles being built starting from a ferrite core, which is then coated with silica gel (greyish), which is in turn studded with gold crystals (little black dots), used as nucleation points to grow the gold shell.

In parallel, a test strip was developed as shown in **Figure 3**, where a sample pad (glass fiber), nitrocellulose, and an absorbent pad were laminated onto a backing card. The key property of the strip was a test line made of a mixture of biotinylated mycolactone analog PG-204 (**Figure 1**) and polystreptavidin, which effectively immobilized the PG-204 probe at the test line. The strip also included a control line of anti-mouse antibody to capture magnetic gold nanoshells that migrated past the test line. The strip was placed in a plastic cassette for ease of use and transportation.

**Figure 3.**
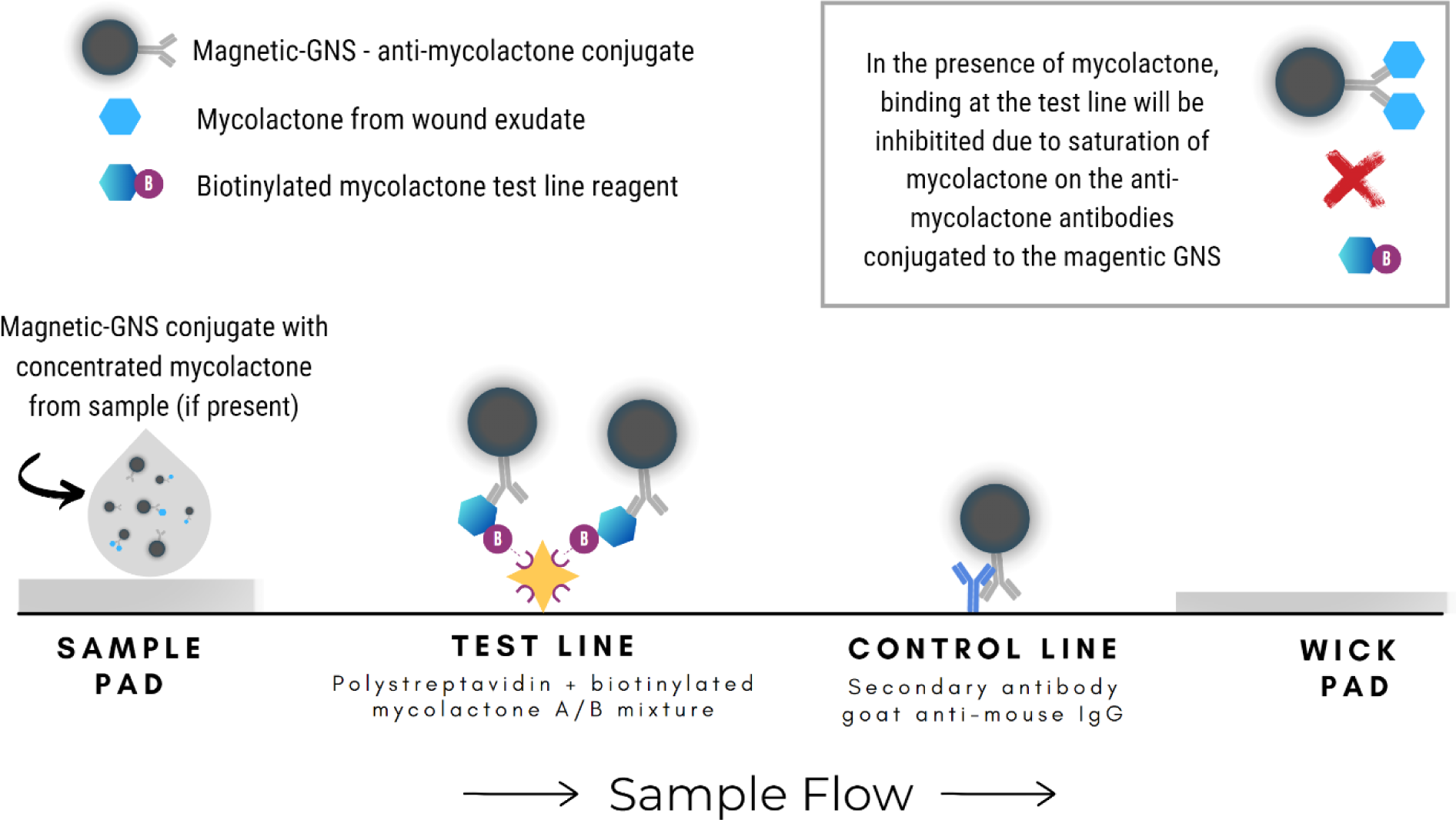
Competitive-format test strip schematic. Biotinylated mycolactone is added to poly-streptavidin and the mixture is immobilized at the test line position. A secondary goat anti-mouse antibody is immobilized at the control line position. The magnetic-GNS anti-mycolactone conjugate will bind to the biotinylated mycolactone at the test line. In the presence of mycolactone in the sample, the mycolactone binding sites on the conjugate will become saturated, resulting in a test line intensity that is inversely proportional to the amount of mycolactone in the sample.

Combined with the magnetic nanoshells covalently conjugated to an anti-mycolactone antibody, the strip allowed for a competitive assay. If the sample to be analyzed is devoid of mycolactone, then the conjugate binds to PG-204, itself immobilized on the test line, and produces a black test line. Conversely, if the test sample contains significant amounts of mycolactone, the latter saturates the binding sites on the anti-mycolactone antibody, and the conjugate is no longer captured by PG-204 at the test line. In summary, the resulting test line intensity is inversely proportional to the concentration of mycolactone in the sample.

### Assay workflow

We envisioned that the workflow of the finished assay would be as shown in **Figure 4**, with its components described in the next paragraphs of the Materials and Methods section. The magnetic nanoshells, after being conjugated to anti-mycolactone antibody (the “conjugate”), would be lyophilized for storage and transportation, and reconstituted in the laboratory or at the point of care. Next, a swab containing a wound exudate would be transferred to a tube containing extraction buffer (step 1). The reconstituted magnetic nanoparticles would be added (step 2), incubated with the extracted sample to capture the mycolactone (step 3), and concentrated by means of a magnetic rack (step 4). The supernatant would be removed (step 5) and the magnetic nanoshells resuspended in chase buffer (step 6) before being added to the lateral flow assay (step 7).

**Figure 4.**
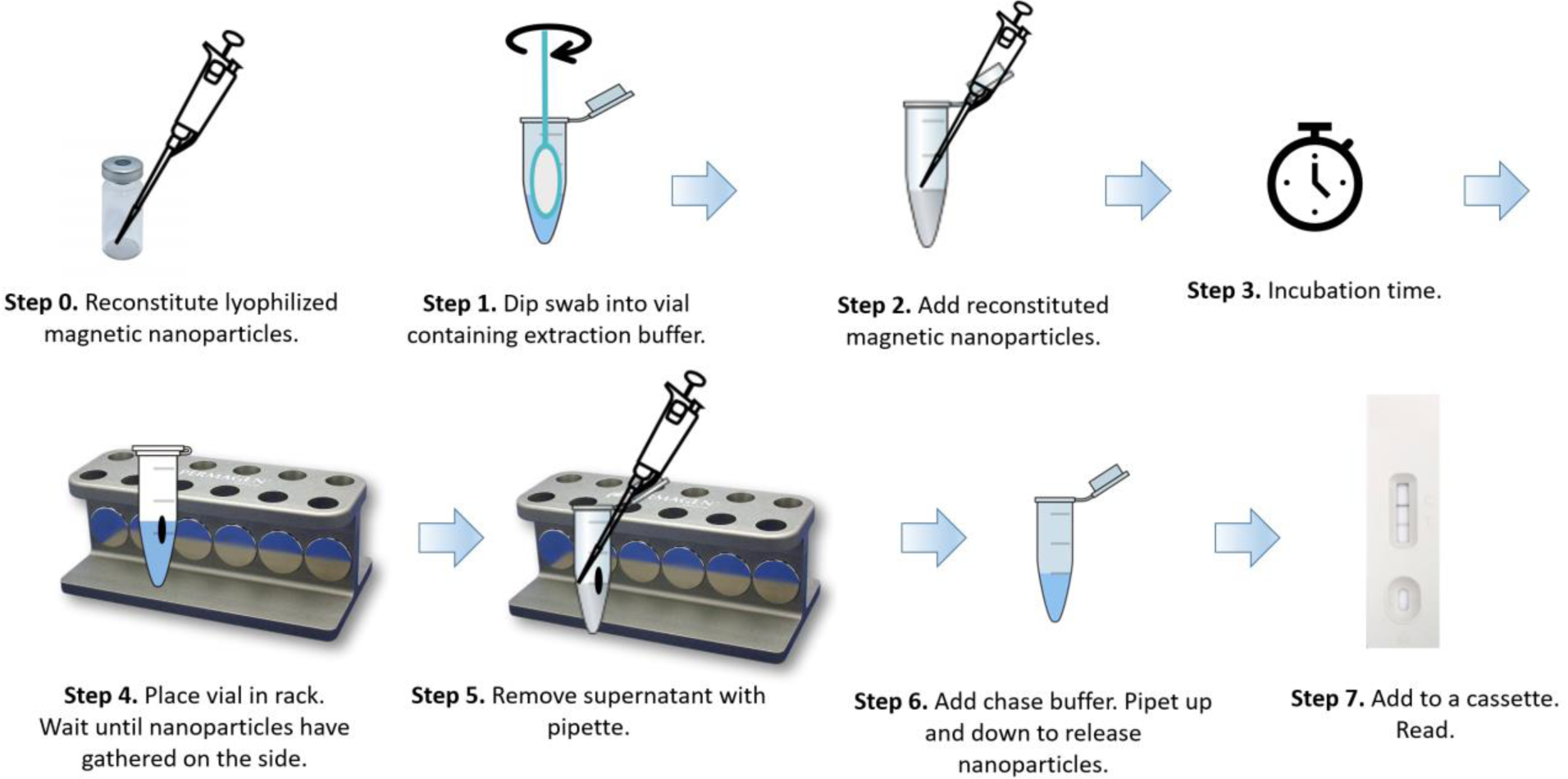
Assay workflow, as initially envisioned.

### Monoclonal antibodies

The generation of mouse monoclonal antibodies to mycolactone has been previously described [17]. The clone chosen for this application was JD5.1, produced by Swiss TPH (Basel, Switzerland) purified from hybridoma culture supernatants by affinity chromatography using a HiTrap Protein A HP column (Cytiva). Two other anti-mycolactone clones were deprioritized: JD5.11 which gave problems with reproducibility, and clone LW2.2 which appeared to have slower binding kinetics.

### Conjugate

Magnetic gold nanoshells were conjugated to mouse monoclonal antibody JD5.1 using EDC/sulfo-NHS chemistry. The magnetic nanoshells conjugated to JD5.1 will be referred to below as “the conjugate”.

### Lyophilization of the conjugate (optional)

The conjugate was stored at 4 °C, and either used as such in solution, or later in the development cycle, in order to match the workflow of **Figure 4**, was freeze-dried as 3% solution in lyophilization medium (5% sucrose, 5% trehalose, 0.1% surfactant 10G in deionized water).

### Mycolactone and mycolactone analogs

Synthetic mycolactone A/B, biotinylated analog PG-204, and acetyl analog PG-119 were prepared by the group of Prof. Altmann, ETH-Z, and stored frozen at –20 °C in DMSO [20, 21]. Mycolactone sourced from WHO in ethyl acetate gave inferior results for reasons that were not investigated and was not further used (data not shown).

### Test strips

The test strips were built using a glass fiber pad (Ahlstrom grade 8951), nitrocellulose type CNPF-SN12, 10 µM (MDI, Advanced Microdevices PVT. LTD) and an absorbent pad (Ahlstrom, grade 222) affixed to a 300 x 60 mm backing card (Lohmann LC-58803) with 2 mm overlaps. The strips were 4 x 60 mm, placed in an off-the-shelf cassette.

### Extraction buffer

Mycolactone was extracted from simulated or real samples using an extraction buffer made of 100 mM triethanolamine (TEA). When testing contrived samples, the pH was adjusted to 7.8. Upon re-optimization with clinical samples, the optimal pH was found to be 10.4. For evaluation of clinical samples, addition of a Heterophilic Blocking Reagent (HBR) was found to be necessary to eliminate matrix effects. For these samples, a heterophilic blocking tube (Scantibodies, cat# PN 3IX762) was reconstituted with 0.5 mL of extraction buffer and used in optimized amounts for each lot of conjugate, at a final concentration of 2–5%. The TEA buffer was stored in glass bottles at room temperature, and HBR was prepared and stored in 1.5 mL Axygen vials (Axygen, cat# MCT-150-L-C) at 4°C.

### Chase buffer

The chase buffer was prepared with 2% proprietary detergent, 0.1% proprietary surfactant, 0.1% casein (Sigma, cat# G7078-500G) prepared by incubation in sodium tetraborate decahydrate pH 8.5 at 37 °C for 24 hours, and 0.05% Proclin (Sigma, cat# 48912-U) in PBS. The pH was adjusted with sodium hydroxide to pH 8.9 for experiments with clinical samples.

### Simulated wound exudate and other test materials

Based on a published biochemical analysis of wound fluids from leg ulcers [22], we concluded that wound fluid could be approximated by mixing equal volumes of negative human serum (NHS, ConeBio, cat# 4090) and PBS (FisherScientific, cat# 28372). We called this 1:1 mixture of NHS:PBS Simulated Wound Exudate (SWE). Positive simulated samples were prepared by spiking SWE with synthetic mycolactone A/B prepared from a mycolactone stock solution (1 mg/mL in DMSO) diluted in ethanol by a factor of 135 to reach a 10 µM concentration. Positive controls were prepared each containing 7 ng of mycolactone per sample, by diluting 1 µL of 10 µM mycolactone in ethanol to a final volume of 75 µL with SWE (for testing without a swab) or by diluting 1 µL of 10 µM mycolactone in ethanol to a final volume of 50 µL with SWE (for testing with a swab). Sterile swabs with a flock-tipped applicator and polystyrene handle (Copan, Cat #25-3306-H BT) were used for testing with swabs.

### Clinical wound exudates

Wound swab extracts from cases that were initially suspected to be Buruli ulcer but later found to be PCR-negative (**Figure 8**) were kindly provided by Dr. Anthony Ablordey from the Noguchi Memorial Institute for Medical Research, University of Ghana. Those extracts were received already extracted in water instead of TEA. Additional samples were collected from suspected Buruli ulcer lesions but that proved negative for mycolactone with our rapid test were provided by The Pasteur Center in Cameroon, Yaoundé, Cameroon, Pasteur Institute Côte d’Ivoire, Abidjan, Côte d’Ivoire, and Hope Commission International, Abidjan, Côte d’Ivoire.

Clinical samples from Ghana were taken at the hospital or health centres as part of the routine diagnostic procedure and case confirmation recommended by WHO and sent to the Noguchi Memorial Institute for Medical Research (NMIMR) for analysis. Ethical approval for collection and use of patients’ samples was obtained from the Institutional Review Board of the Noguchi Memorial Institute for Medical Research (NMIMR; Study number: NMIMR-IRB CPN 024/18–19). Patients were required to sign an informed consent. To ensure anonymity, personal information such as names, and other personal identifiers associated with patient’s samples were replaced with codes. Similarly, Clinical Samples from Ivory Coast and Cameroon were collected under IRB approved protocols for the sake of this study and with informed consent. The protocols were cleared by the Comité National d’Ethique des Sciences de la Vie et de la Santé, Ministère de la Santé, Côte d’Ivoire, 2021, IRB 000111917 and the Comité National d’Ethique de La Recherche pour la Santé Humaine, Ministère de la Santé, Cameroun, IRB 2021/06/1367.

### Mycobacteria cultures

*Mycobacterium marinum* and *Mycobacterium ulcerans* were cultured as previously described [23].

### Procedure to run the assay using a simulated sample and no swabs (**Figure 4** Steps, 2-7)

For the sake of simplicity, most of the assay development work was carried out without a swab, skipping steps 0 and 1 of Figure 4. Instead, 75 µL of negative SWE or positive (mycolactone spiked) SWE were placed in a 1.5 mL Axygen vial (Axygen, cat# MCT-150-NC) and diluted with 0.5 mL of extraction buffer. Then 1–2 µL of conjugate Mag-GNS-JD5.1 in solution (OD= 20, or an equivalent of the reconstituted lyophilized conjugate) was added to the extract, vortexed briefly, and incubated for 30 min, then placed on a magnetic rack to concentrate the nanoparticles for 30 min. During incubations the vials were protected from the light to avoid mycolactone degradation. The conjugate was concentrated with a neodymium magnet (Poichekailov magnetic racks, US, [24]), the supernatant was removed and the particles resuspended in 2 drops (approximately 60 µL) of chase buffer and transferred to the test sample port. The test developed for 30 min prior to imaging and quantitation with a benchtop reader (Leelu v3.0, Lumos Diagnostics, Carlsbad, CA).

### Procedure to run the assay using a simulated sample absorbed onto a swab (**Figure 4**, Steps 1-7)

To simulate a clinical specimen absorbed on a swab, a swab (Copan, 25-3306-H BT) was placed in an Axygen vial containing 50 µL of positive or negative SWE until all the liquid was absorbed. Next, the swab was placed in a fresh 1.5 mL Axygen vial and extracted with 0.5 mL of extraction buffer. The swab was rotated 3–5 times in the extraction buffer while pressing against the sides of the vial, left in the vial for 5 min, and pressed against the wall while rotating and lifting to recover as much liquid as possible. The swab was discarded, and the next steps (steps 2–7) were conducted with the extract as described in the previous paragraph for SWE.

### Procedure to run the assay with clinical wound exudates (with HBR, pH 10.4 buffer)

The retrospective PCR negative clinical wound swabs from Ghana had been extracted in water prior to receipt. These samples were tested with two methods (1) as received, following the protocol for simulated extracts and (2) by first adding 10x concentrated TEA extraction buffer to final x1 concentration to improve conjugate collection on a magnet. A heterophilic blocking reagent (HBR) was added to clinical wound extracts at 2– 5% concentration to prevent possible interference of the conjugate with heterophilic antibodies or other matrix effects, then samples were tested the same as described for the simulated extract.

### Quantification

Test line intensities were quantified with a benchtop reader (Leelu v3.0, Lumos Diagnostics, Carlsbad). We investigated both the test line intensity alone, as well as the ratio between test and control line (T/C) with the latter giving the most reproducible results.

### Statistical Analysis

P-values were calculated using GraphPad Prism, version 10, assuming Gaussian distributions, and not assuming equal standard deviations (Welch’s correction).

## Results

### Optimal conditions

As described under Materials and Methods, based on limited literature [12] our initial estimate was that clinically relevant concentrations of mycolactone would be 15–150 ng/swab. Opting for a conservative approach, we aimed at developing a test that could detect half of the lowest amount that we were expecting to find in swabs (*i.e.,* approximately 7.5 ng of mycolactone). To standardize the testing method and to overcome the paucity of clinical materials, we prepared contrived samples in which wound exudate was simulated by using a 1:1 mixture of negative human serum and PBS. The simulated exudate was then spiked with known amounts of synthetic mycolactone A/B.

Assay optimization was extensive and is not described herein. Optimization was first performed with simulated wound exudates, without employing a swab. With this initial set of optimized conditions, the contrived sample (50–75 µL) was diluted in extraction buffer (0.5 mL, pH 7.8), and incubated with the magnetic conjugate (1–2 µL, OD 20) for 30 minutes. The magnetic conjugate was then recovered by placing it on a magnetic rack for 30 minutes, removing the supernatant, and adding 2 drops (approximately 60 µL) of chase buffer (pH = 7.4). The resuspended nanoparticles were then added to the sample port of the cassette, and the assay ran for 30 minutes before analysis.

These conditions worked well with contrived samples but as described below, failed when using clinical samples. This triggered a second round of optimization, which led to increasing the pH of the extractions and chase buffers while leaving the other parameters unchanged. Thus, when using clinical samples, we employed a TEA extraction buffer adjusted to pH = 10.4 instead of pH = 7.8 (0.5 mL). The incubation with the magnetic conjugate (1–2 µL, OD 20, 30 min) remained the same, and so did the magnetic conjugate recovery method (magnetic rack, 30 min, and removing the supernatant). The newly optimized chase buffer was adjusted to a pH = 8.9 instead of pH = 7.4 (2 drops). The rest of the procedure was not modified: the resuspended nanoparticles were then added to the sample port of the cassette, after which the test developed over 30 minutes. The fully optimized conditions can be found in the Instructions for Use included in the Supplementary Information.

### Limit of detection with simulated wound exudate

Samples of mycolactone A/B serially diluted in ethanol were added to the simulated wound exudates and processed according to the optimized conditions described in the previous two paragraphs. At this stage of development, the extraction buffer had a pH = 7.8 and the chase buffer had a pH = 7.4. The intensity of the test and control lines were quantified with a benchtop reader. The more mycolactone in the sample, the more it competes with PG-204 immobilized at the test for binding the conjugate. Hence, the higher the mycolactone concentration, the lower the test line intensity. When looking at the test with the unaided eye (**Figure 5**), the test line could be easily seen if no mycolactone was present, and the test line became barely visible when 7 mg of mycolactone were added to the simulated wound exudate. Similarly, when quantifying the intensity of the test lines with a benchtop reader (**Figure 6**), the intensity decreased from approximately 0.08 reader units in the absence of mycolactone to 0.02 units in the presence of 7 ng mycolactone, and the difference was statistically significant (p=0.006). All samples contained the same amount of DMSO, indicating that the dose response was due to mycolactone and was not an artifact due to DMSO.

**Figure 5.**
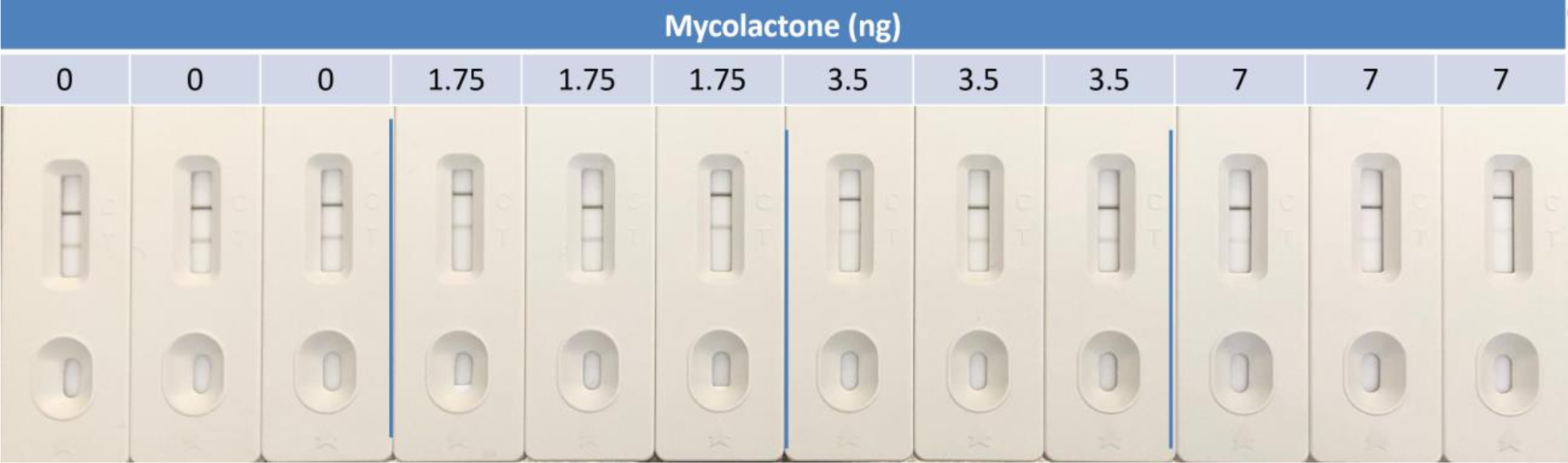
Limit of detection of the rapid test using simulated samples spiked with synthetic mycolactone (n=3). Visually, a difference can be seen between no mycolactone and 7 ng of mycolactone, where the latter causes a nearly complete inhibition of the test line.

**Figure 6.**
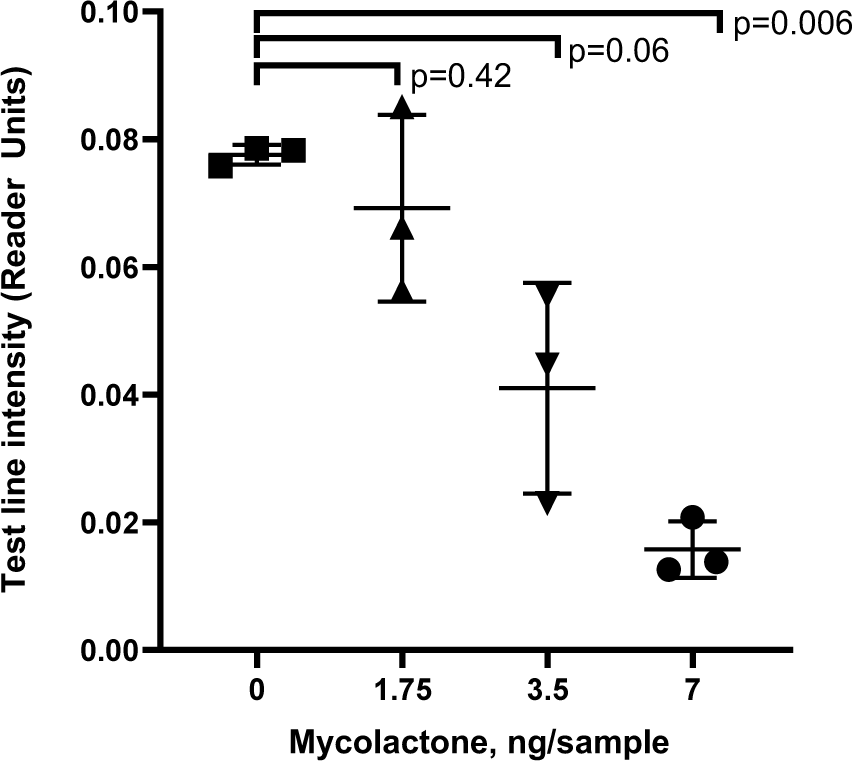
Limit of detection of the rapid test using simulated samples spiked with synthetic mycolactone. When measured with a reader, the difference between no mycolactone and 7 ng of mycolactone is statistically significant (p < 0.05).

### Specificity with cell cultures of different mycobacteria strains

Having developed a test that appeared to have the desired analytical sensitivity, we next evaluated its specificity. This was done by comparing supernatants from laboratory-grown cell cultures of *M. ulcerans* and *M. marinum,* a non-pathogenic mycobacterium lacking the giant 174 kb symbiotic plasmid pMUM001 responsible for mycolactone production in *M. ulcerans.* As shown in **Figure 7**, *M. ulcerans* supernatant, which contains mycolactone, led to an inhibition of the test line, while *M. marinum* cell cultures, which do not contain mycolactone, did not inhibit the test line formation, demonstrating that our assay shows no cross-reactivity with *M. marinum* samples.

**Figure 7.**
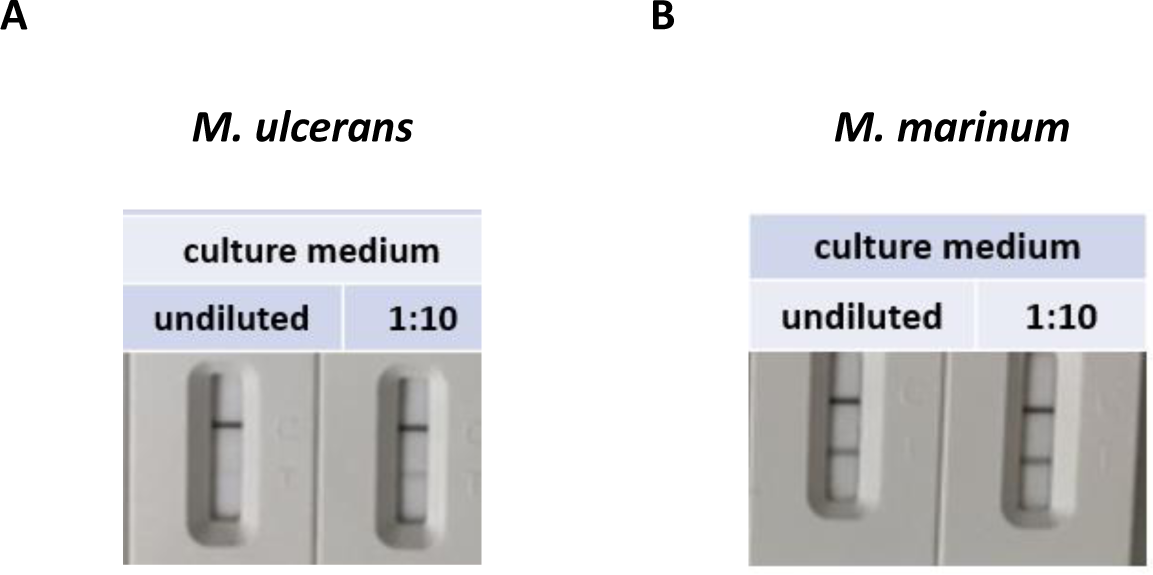
Cross-reactivity testing the assay with *M. ulcerans* vs *M. marinum* supernatant. While *M. ulcerans* produces mycolactone leading to an inhibited signal at the test line, *M. marinum* does not synthesize mycolactone and therefore the test line remains visible.

### Test results with swabs and simulated wound exudates

Having in hand a test that was analytically sensitive and specific, we next investigated the use of a swab. The concern was that a swab would be mandatory for sample collection but that mycolactone, being so lipophilic with a calculated logP of 9.0 [25], may stick to the fibers of which the swab is made, and not be easily extracted into an aqueous solution. To investigate this matter, an aliquot of 50 µL SWE was deposited on a swab and then extracted with the extraction buffer for 5–10 min. As reference, the same SWE was directly diluted with extraction buffer, without using a swab. The tests showed a nearly identical behavior when directly diluting the SWE in extraction buffer (not using a swab) versus first absorbing the SWE onto a swab and then extracting it (**Figure 8A**). Based on the quantification of the test line intensities, we calculated that 80% of the mycolactone was recovered from the swab, and the remaining 20% were left within the extract trapped in the swab fibers. Additionally, the swabs containing simulated samples could be stored for at least 48 hours at 4 °C before extraction without noticeable degradation of mycolactone (**Figure 8B**). This feature is useful for shipping samples from the field to a local laboratory.

**Figure 8.**
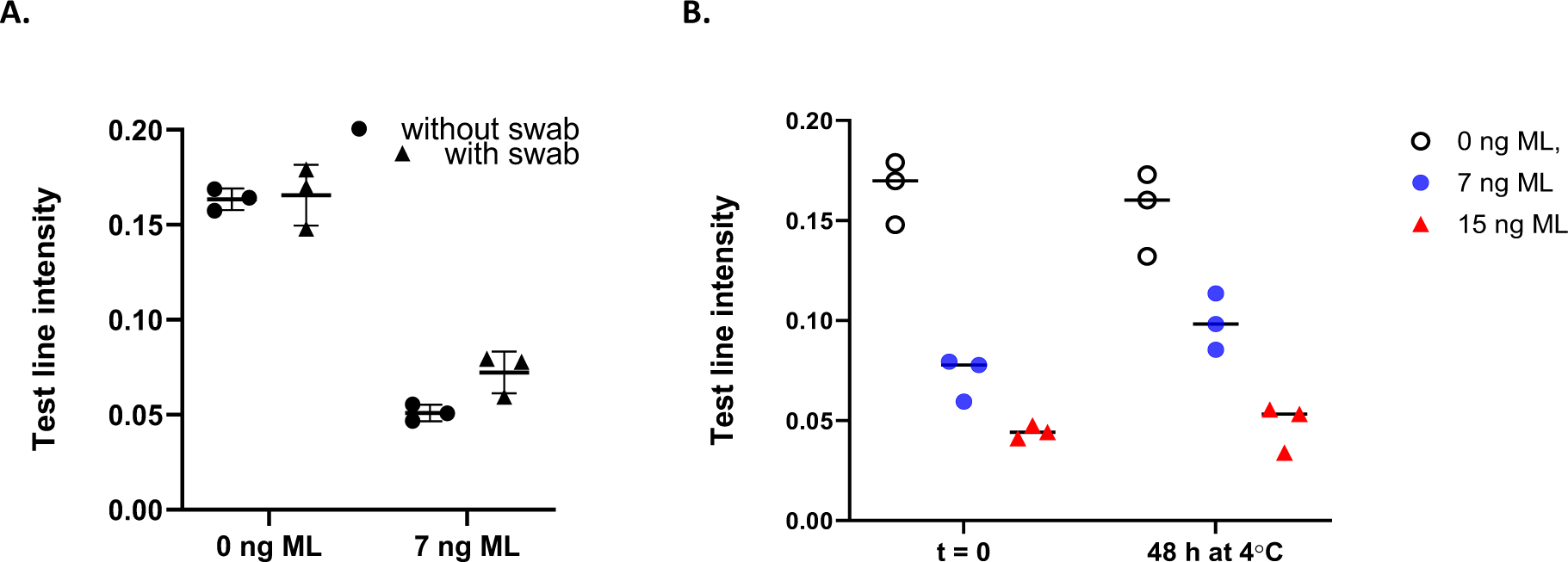
A. Effect of swab on test results and sample stability when absorbed on a swab. Panel A. The test results are not impacted if simulated wound exudate is pre-absorbed on a swab. Panel B. Once absorbed on a swab, simulated wound exudates are stable for at least 48 hours at 4 °C.

### Optimizing the test conditions to real wound exudates

Having established the performance of the test with simulated wound exudates, we moved to real wound exudates, collected from ulcers in Buruli ulcer-endemic areas (Ghana). Contrary to simulated wound exudates, real clinical wound exudates collected in the field represented a challenging matrix that varied from patient-to-patient and included blood and various other debris (**Figure 9**).

**Figure 9.**
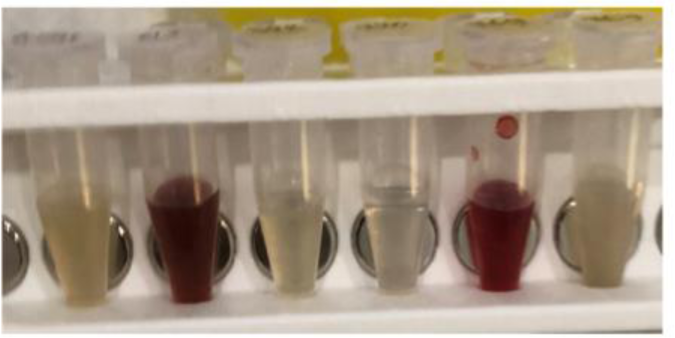
Aspect of 6 different wound exudate extracts as received from Ghana.

To our surprise, both the test and control line frequently disappeared, even though the samples had tested negative for Buruli Ulcer by PCR. Some clinical samples contained debris and particulates, sometimes in large amounts and possibly deriving from herbal medications and necrotic tissues. These debris interfered with the test, although could not explain alone all the cases where test and control lines disappeared.

Several parameters were optimized to achieve 100% validity (*i.e.,* control line always visible) with clinical swabs. First, we reasoned that the test employs mouse monoclonal antibodies, and that heterophilic antibodies present in the wound exudates may have a deleterious impact, especially when using endemic samples from populations often exposed to rodents. This risk was mitigated by adding Heterophilic-Blocking Reagents (HBR) which led to a significant improvement of the line intensities (**Figure 10**).

**Figure 10.**
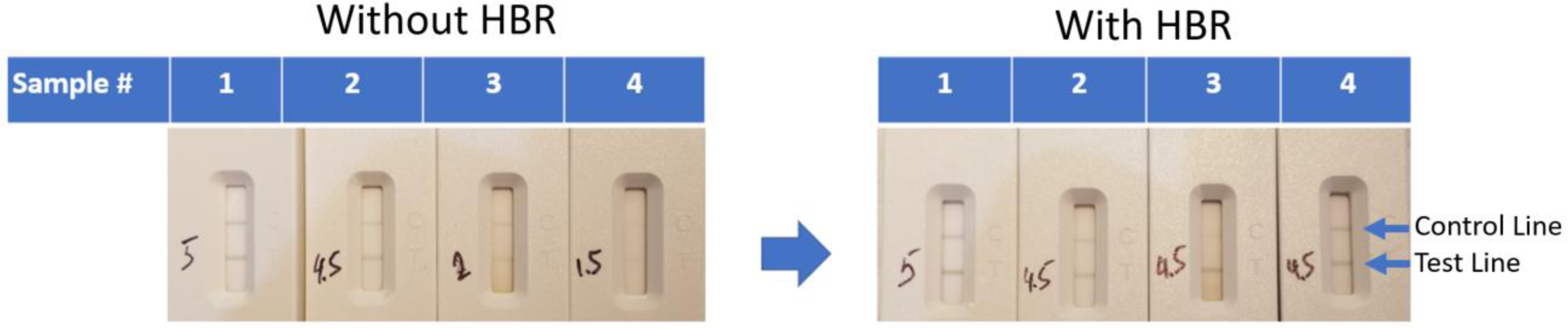
HBR rescues the control line with negative wound swabs and improves the aspect of all tests. The test line intensity determined with a visual score card is indicated on the cassettes next to the test line.

Second, we removed the particulate material from the swab extracts by centrifugation. This can be done easily in the laboratory but not in the field. Alternatively, doubling the amount of buffer with which the mycolactone is extracted from the swab (1.0 mL instead of 0.5 mL) lowered the impact of particulate on magnetic nanoparticle recovery (data not shown). Third, increasing the pH of both the extraction buffer (from 7.8 to 10.4) and chase buffer (from 7.4 to 8.9) increased the intensity of both the control and test lines (**Figure 11**). Taken together, these modifications provided 100% validity, as judged by the presence of a visible control line in all of the 207 clinical swabs analyzed.

**Figure 11.**
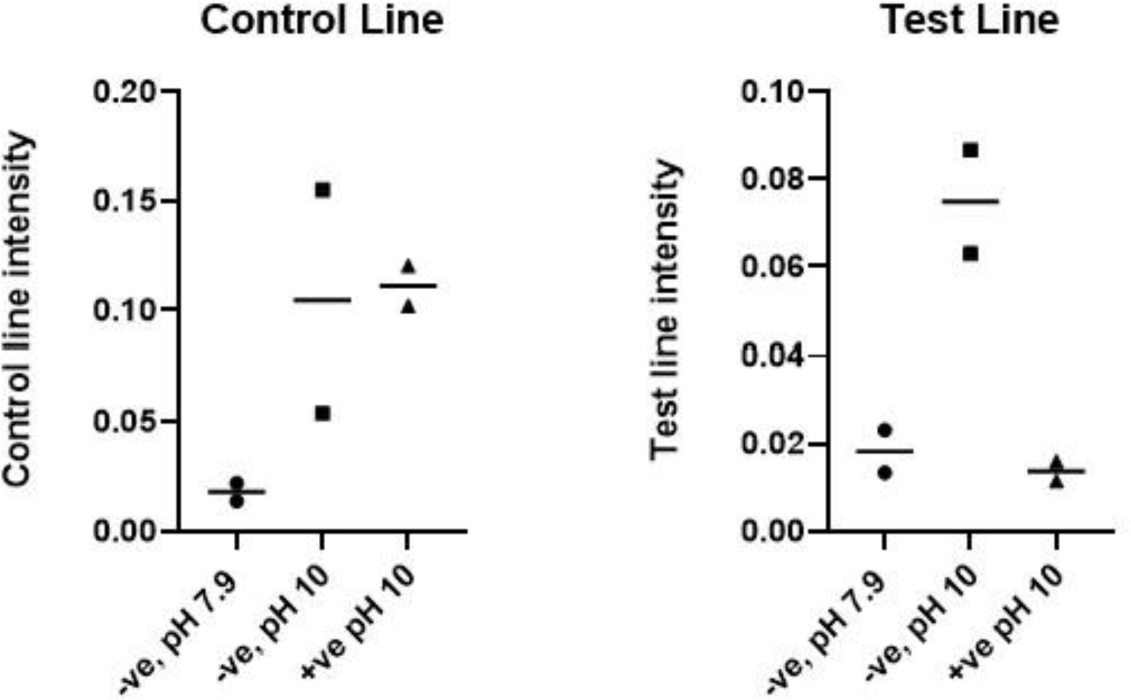
Increasing the pH of the extraction buffer from 7.9 to 10 leads to stronger control lines (left panel) and test line intensity (right panel) when testing simulated wound exudate samples, either not spiked with mycolactone (negative; -ve) or spiked with 7 ng mycolactone (positive; +ve).

### Calibration curve and limit of detection (LOD) with real wound exudates

After optimizations (sample centrifugation, addition of HBR and pH optimization of extraction and chase buffers) were introduced we re-examined the dose-response curve using real exudates extracted with TEA pH 10.4. Wound extracts from LFA-negative lesions were pooled, and then spiked with synthetic mycolactone. Rather than reporting the absolute intensity of the test line as we did in the original LOD testing (**Figures 5, 6**), the test to control (T/C) line ratios were calculated, leading to more consistent results (**Figure 12**). In the absence of mycolactone, the T/C ratio was around 0.9. The presence of 3.7 ng of mycolactone/sample gave a T/C ratio in the 0.42–0.44 range that was easily distinguished from the T/C ratio obtained without mycolactone. Hence, under fully optimized conditions, the LOD was around 3.7 ng of mycolactone using extracts from clinical samples as the sample matrix.

**Figure 12.**
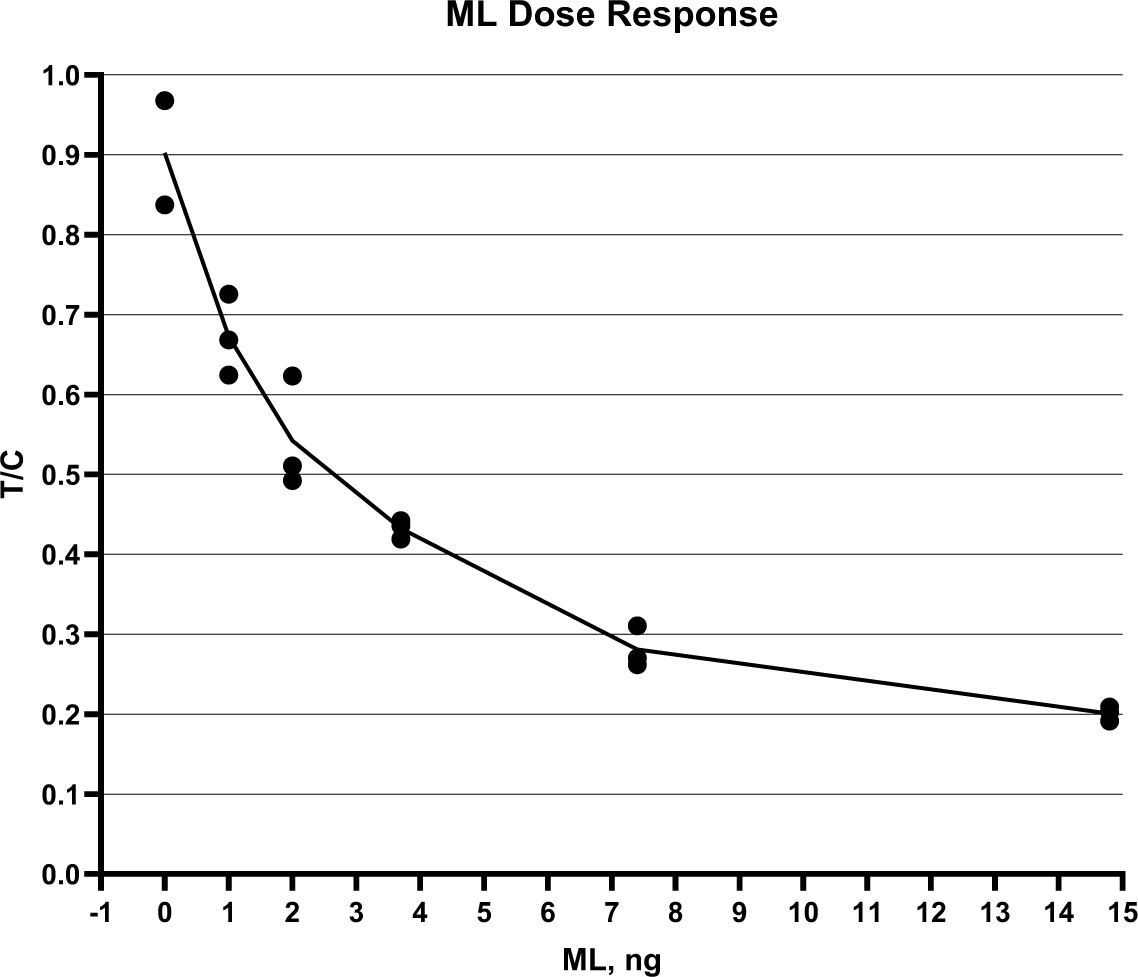
Mycolactone dose response in pooled Buruli ulcer negative swab extracts from Cameroon spiked with synthetic mycolactone A/B. T/C – test-to-control line ratio quantified with Lumos Leelu benchtop reader.

### Additional work

Additional work that is not described herein included the freeze-drying of the conjugate, and the demonstration that once lyophilized the conjugate is stable for at least 2 weeks at 40⁰C, allowing for it to be shipped to disease endemic countries and field settings without prohibitive cold-chain requirements. We also demonstrated that a positive control could be made with mycolactone analog PG-119 (**Figure 1**). Analog PG-119, which lacks the side-chain of mycolactone, is also detected by monoclonal antibody JD5.1, with the same or better sensitivity as mycolactone A/B. This offers two major advantages, as PG-119 is easier to synthesize and is not photosensitive.

## Discussion

### Target product profile

Based on the WHO target product profile (TPP) for Buruli ulcer, we developed a rapid diagnostic tests (RDT) targeting mycolactone. While the TPP specifies mycolactone as the analyte of choice, it does not specify the required limit of detection. Based on very limited literature data [12, 26], we inferred that a test capable of detecting 15 ng of mycolactone would provide the required clinical sensitivity. To be conservative, we defined that the limit of detection (LOD) should be 7 ng of mycolactone per sample or lower. It remains to be seen if this truly translates to proper clinical sensitivity [27].

Developing a mycolactone-based test was difficult for several reasons. Firstly, the matrix in which mycolactone is found is complex. The disease starts as a papule, that turns into a nodule, and eventually into a large open wound (ulcer). Nodules can be sampled by means of fine needle aspirates, while wounds are sampled with swabs. Since people are more likely to seek treatment after the appearance of an ulcer, we elected to detect mycolactone in wound exudates collected with swabs. Wound exudates are an unusual matrix for lateral flow assay development, as they vary in physical appearance and composition from one person to another – for instance some lesions can be relatively dry and others exudative. Another limitation of wound exudates is that they are available only in small amounts, unlike more mainstream matrices such as plasma or urine. Furthermore, the amount of mycolactone in wound exudates can only be limited, in part because *M. ulcerans* replicates slowly, therefore generating only low concentrations of mycolactone, and because mycolactone is photodegradable [28] and hence can be cleared from the wound. Another complication is that it is not always clear if patients have applied herbal medications or other topical treatments to the ulcer and how that may affect the viability of *M. ulcerans* and its ability to produce mycolactone, and if those treatments have potential to interfere with the test. Finally, mycolactone is a highly hydrophobic molecule (cLogP = 9.0) [29, 30], and therefore has low affinity for aqueous mediums and can stick to laboratory plasticware.

The developed Buruli ulcer RDT was configured as a competitive assay to detect mycolactone using novel magnetic gold nanoshells, mycolactone-specific monoclonal antibodies [17] and a biotinylated probe. Unlike normal RDTs, a mycolactone concentration step was required. This was accomplished by leveraging the magnetic properties of the nanoparticles, allowing the assay to be performed in under two hours, with minimal equipment (**Figure 1**). We initially noticed a strong difference between simulated wound exudate, prepared by mixing equal volumes of negative human serum and PBS, which gives clean test results, and real wound exudates which gave in some cases invalid tests, with no test line and no control line. In these problematic cases, the nanoparticles seemed to be especially difficult to recover magnetically. This behavior was tentatively ascribed to the presence of heterophilic antibodies recognizing the mouse anti-mycolactone antibody, and either modifying the charge of the surface of the gold nanoparticle or causing the nanoparticles to cross-link with each other and complicating their recovery. Regardless of whether this rationale is correct or not, the unwanted absence of control line could be corrected by adding HBR to the extraction buffer and by using higher pH in the extraction and chase buffers. The addition of HBR added an extra step to the workflow presented in Figure 1, and the final workflow can be found in Supplementary Information section (**Figure S1 and Instructions for Use**).

The limit of detection of the assay was 3.7–7 ng mycolactone per sample analyzed, and this was estimated to be sufficient for the test to be clinically relevant. We verified that the magnetic nanoparticles could be freeze-dried and be stable in this format at 40 °C for at least 2 weeks for shipping purposes. We also verified that mycolactone is stable for at least 2 days at 4 °C once absorbed on a swab, enabling the samples to be transported from the field to a district laboratory. The materials (swabs, vials) selected in this publication were compatible with mycolactone and allowed for maximum recovery.

In conclusion, a prototype RDT was developed that met the WHO TPP specification of detecting mycolactone as the biomarker, and that met the self-imposed requirement of detecting at least 7 ng of mycolactone in wound exudate. The test also met most of the WHO TPP parameters in that it was stable for transportation at ambient temperature, and that a stable positive control was developed. Detailed instructions for use were compiled and are available in the Supplementary Information. Based on this data, the prototype was deemed worthy of advancing to field evaluations, with fresh clinical samples. These results will be reported elsewhere.

## Supporting Information

- Instructions for Use (pdf)
- Finalized workflow (Figure S1)
- Aspect of nanoparticles on the rack (Figure S2)

## Data Availability

All relevant data is included in the manuscript

## Acknowledgements

This work received financial support from FIND through funding from Medicor Foundation, UBS Optimus Foundation, Swiss Agency for Development and Cooperation (SDC), KfW-BMBF and Global Health Innovative Technology (GHIT) Fund. The funders played no role in the decision to publish. The authors thank all those involved in sample collection and field testing of the device, including Dr. Sara Eyangoh, Mr. Hycenth Numfor, and Ms. Valerie Donkeng Donfack and the team from Buruli Ulcer Laboratory Network (BU-LABNET), Centre Pasteur du Cameroun, Dr. Aka N’guetta and Dr. David Coulibaly from the Institute Pasteur de Côte d’Ivoire, Mr. Aubin Yao from the Hope Commission International, Côte d’Ivoire and the team from the National Buruli Ulcer Control Program of Côte d’Ivoire. And Dr. A. Ablordey and his team at the Noguchi Memorial Institute for Medical Research.

